# Clinical Characteristics and Outcomes of COVID-19 Positive Acute Coronary Syndrome Patients; a multisource Electronic Healthcare Records Study from England

**DOI:** 10.1101/2020.08.20.20175091

**Authors:** Muhammad Rashid, Jianhua Wu, Adam Timmis, Nick Curzen, Sarah Clarke, Azfar Zaman, James Nolan, Ahmad Shoaib, Mohamed O Mohamed, Mark A de Belder, John Deanfield, Chris P Gale, Mamas A. Mamas

## Abstract

**Background:** Patients with underlying cardiovascular disease and Coronavirus disease 2019 (COVID-19) infection are at increased risk of morbidity and mortality. However, there is limited information on management and outcomes of patients presenting with acute coronary syndrome (ACS) and concomitant COVID19 infection.

**Objectives:** This multisource national analysis of live data from England was designed to characterise the presenting profile and outcomes of patients hospitalized with ACS and COVID-19 infection.

**Methods:** Multisource data from all acute NHS hospital in England was linked to study the characteristics and outcomes of patients hospitalized with COVID-19 ACS compared to non COVID-19 ACS patients. Hierarchical multilevel models were constructed to study the association between COVID19 ACS and in-hospital and 30-day mortality.

**Results:** Between 1st March 2020 and 31st May 2020, 517 (4.0%) were admitted with COVID-19 ACS from a total of 12,958 ACS patients. COVID-19 ACS patients were generally older, BAME ethnicity, more comorbid and had unfavourable presenting characteristics compared to non-COVID-19 ACS patients. They were less likely to receive invasive coronary strategy in the form of coronary angiography (67.7% vs 81.0%), PCI (30.2% vs 53.9%), dual antiplatelet medication 76.3% vs 88.0%), and other important secondary medication. Patients with COVID-19 ACS had higher in-hospital (aOR 3.27 95%CI 2.41-4.42) and 30-day mortality (aOR 6.53 95%CI 5.1-8.36) compared to non COVID-19 ACS group.

**Conclusion:** COVID-19 infection is prevalent but less frequent in the patients hospitalized with ACS in England. Presence of COVID-19 infection in patients with ACS is associated with significant mortality hazard.

## Introduction

Coronavirus disease 2019 (COVID-19), caused by severe acute respiratory syndrome coronavirus-2 (SARS-CoV-2), has affected more than 17 million people resulting in almost 700,000 deaths worldwide^1^. Although, COVID-19 patients predominantly present with respiratory symptoms, various extra-pulmonary manifestations including thrombotic events, myocardial injury and ischaemia, acute kidney injury and cardiac arrhythmias have also been reported^2, 3^.

Acute coronary syndrome (ACS) in the context of viral infection may be related to atherosclerotic plaque rupture precipitated by endothelial cell damage, a cytokine storm and a heightened inflammatory state^4^ Furthermore, admission with an ACS during the COVID-19 pandemic in which large numbers of patients were hospitalised with COVID-19, may increase the risk of nosocomial transmission of COVID-19 in this vulnerable patient group.

The management of patients presenting with suspected ACS in the context of COVID19 remains a challenge^7^. There are limited data regarding the clinical characteristics, management strategies and post-discharge mortality of patients hospitalized with a diagnosis of ACS and concomitant COVID-19 infection^7^. Small case series of 18 and 28 patients presenting with ST-elevation acute myocardial infarction (STEMI) and concomitant COVID-19 infection have reported significant variability in presenting characteristics and in-hospital survival of these patients^8, 9^. These reports lacked data around clinical presentation, pharmacological treatments and post-discharge survival. Fewer data are available from a national, or from a broader ACS perspective including non-ST-elevation myocardial infraction (NSTEMI). Such information could prove useful in order to devise optimal pathways of care in the event of a second wave of COVID-19.

This study, using high resolution, multisource contemporary national data from England, systematically profiles the presenting and procedural characteristics, in-hospital and 30-day mortality in patients admitted with a suspected diagnosis of ACS and concomitant COVID-19 infection. The primary aim was to investigate the in-hospital and 30-day all-cause mortality in patients hospitalized with COVID-19 ACS diagnosis compared with those without COVID-19 ACS. The second aim was to describe the differences in management and independent predictors of 30-day mortality of those with COVID-19 ACS.

## Methods

### Study data

An unselected, real-world cohort of all patients hospitalized with a suspected diagnosis of ACS in England was derived by linking patient records across four different sources of data, namely Hospital Episode Statistics (HES), the Myocardial Ischaemia National Audit Project (MINAP), the British Cardiovascular Intervention Society (BCIS) percutaneous coronary intervention registry and Civil Registration Death data. The full details about the validity, strengths, limitations and utility for research purposes have been described previously^10–12^. Briefly, HES contains International Statistical Classification of Disease-10^th^ Revision (ICD-10) clinical, geographic, administrative and patient information of all patients hospitalized in any National Health Service (NHS) hospital in England^10^. MINAP is an exclusive ACS registry designed to collect information across 130 data fields about patient demographics, use of various pharmacological and invasive treatments and in-hospital care of patients hospitalized with a suspected diagnosis of ACS (type 1 myocardial infarction) in any NHS acute care hospital in England^12^. Similarly, almost over 98% of the PCI activity in England is captured in the BCIS PCI registry, which is designed to collect detailed procedural and clinical data of all patients undergoing PCI^11^. Finally, the civil registration of death register holds the mortality information of all deaths in England. All patient records in these datasets can be identified using a unique 10-digit number. For the purposes of this study we used live reporting data from all hospitals in England submitting their data to these respective registries during the COVID-19 pandemic.

### Ethical approval

This study was conducted under the endorsement of the Chief Scientific Advisor to the UK government and Scientific Advisory Group for Emergencies (SAGE). The access to the data was granted under the Health and Social Care State Secretary’s notice issued under Regulation 3(4) of the NHS (Control of Patient Information Regulations) 2002 (COPI) to NHS Digital, allowing NHS Digital to share confidential patient information with organisations entitled to process this under COPI for COVID-19 purposes. Furthermore, MINAP and BCIS data are collected and hosted by the National Institute of Cardiovascular Research (NICOR) and used for audit and research purposes without formal individual patient consent under section 251 of the NHS Act 2006^13–16^. The study complies with the Declaration of Helsinki.

### Study cohort

The analytical cohort for this study consisted of consecutive patients aged ≥18 years hospitalized with a suspected diagnosis of ACS in England and documented within the MINAP registry, between 1^st^ March 2020 to 31^st^ May 2020. The COVID-19 status information for all patients was derived from HES using ICD-10 codes ‘U071’ (confirmed) and ‘U072’ (suspected) and linked with the MINAP record of the same patient matching across the two datasets based on their unique NHS number and admission date. In the second step, the records of all these patients were linked across to the records in the BCIS PCI registry using the same NHS number and admission week. Finally, the mortality information of all patients was tracked within 30 days beyond discharge or up to 10^th^ July 2020 from the Civil registration of Death register. Supplementary figure 1 illustrates the cohort selection and data linkage steps of all patients in the study.

**Figure 1:**
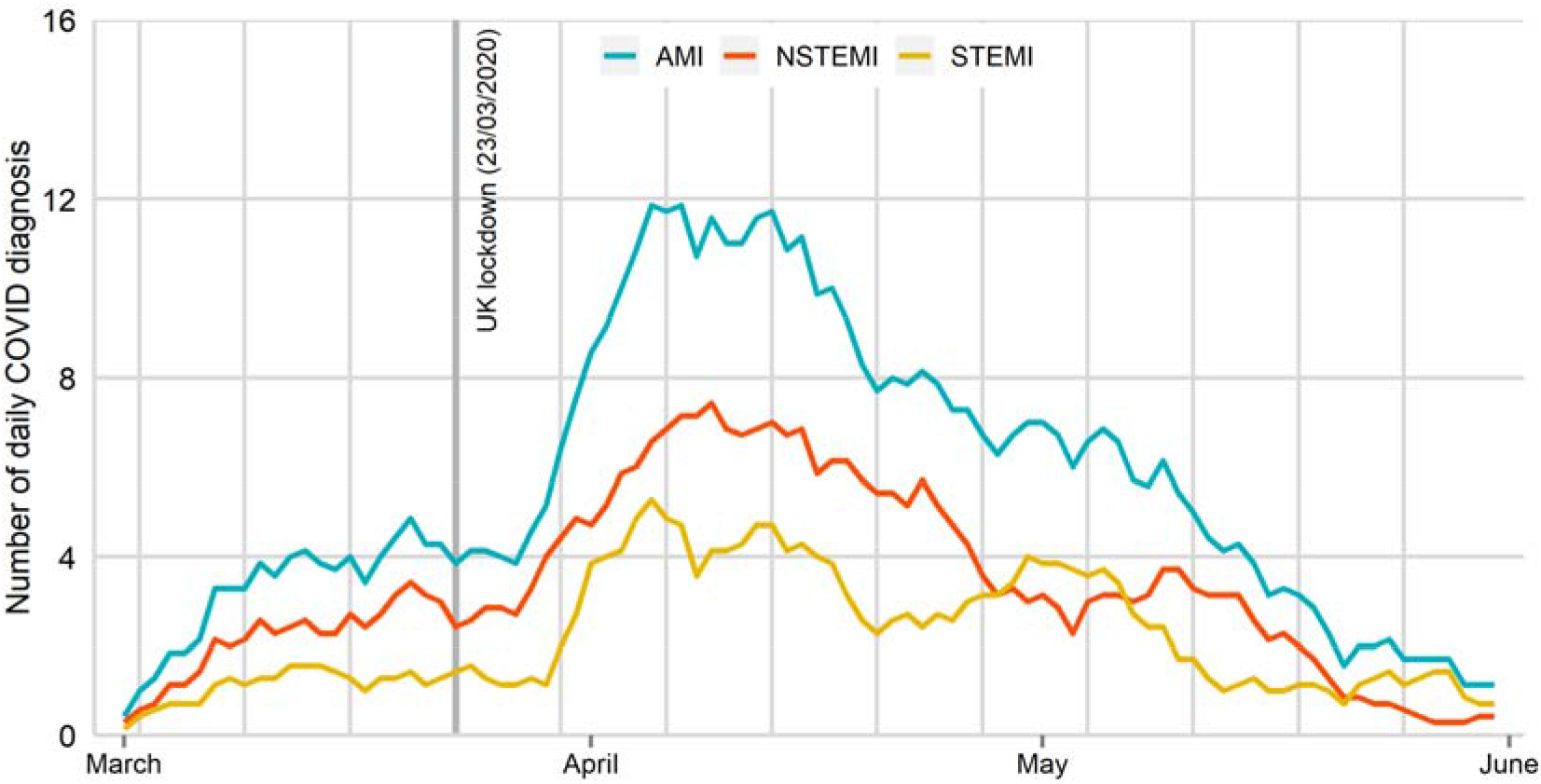
Daily cases of COVID19 ACS hospitalized during the study period

Final study data contained comprehensive detailed information about patient demographics, COVID-19 status, clinical characteristics, comorbidities, pre-hospital and in-hospital pharmacological treatments, cardiac investigations, invasive coronary procedures, procedural characteristics and complications. Patients with missing or invalid NHS number and date of admission were excluded from the analysis. Patients with valid diagnosis codes of COVID-19 captured in the HES data were defined as the “COVID-19 ACS” group whilst all other ACS patients were defined as the “non COVID-19 ACS” group. Time to reperfusion therapy for STEMI was calculated from time of admission to time of receipt of reperfusion treatment where time of admission was defined as time of arrival into hospital. The primary outcomes of interest were in-hospital and 30-day mortality in ‘COVID-19 ACS’ compared with ‘non COVID-19 ACS’ patients.

### Statistical analysis

Baseline characteristics were reported as number and percentages for categorical variables, means with standard deviation or median with interquartile ranges for continuous data. Statistical differences between the two groups were obtained using Chi-Square test or t-test or Kruskal-Wallis tests as appropriate. The daily counts of COVID-19 positive ACS patients were presented using a simple daily moving average from 1st March 2020 up to and including 31^st^ May 2020, adjusted for seasonality. Multiple imputations using chained equation (MICE) techniques were used to impute missing values in all the study variables. Each of the imputation models included all the other variables used in the analyses as reported in the online supplement (Supplementary Table 1). Linear regression for continuous variables, multinomial for nominal variables, ordinal logistic regression for ordered factors and logistic regression models for binary variables were used to generate 10 imputed datasets and all subsequent analyses were performed on these and results were pooled using Robin’s rule^17, 18^.

To study the association between ‘COVID-19 ACS’ status and clinical outcomes, hierarchical logistic regression models with random intercept were constructed. The hospital ID was used as a random intercept to account for nesting of patients within the different hospitals. All models were adjusted for age, sex, ethnicity, body mass index, presenting characteristic (blood pressure, heart rate, cardiac arrest, clinical syndrome, creatinine, Kilip class, left ventricular systolic function), cardiovascular comorbidities (previous PCI, previous CABG, previous AMI, previous cerebrovascular event, peripheral vascular disease, renal dysfunction, heart failure, hypercholesterolemia, angina, diabetes, smoking status, asthma or chronic obstructive airway disease and family history of coronary heart disease), in-hospital pharmacology (low molecular weight heparin, unfractionated heparin, warfarin, loop diuretic, glycoprotein IIb/IIIa inhibitor), discharge pharmacology (dual antiplatelet medication use, statin, angiotensin converting enzyme (ACE) or angiotensin receptor blocker use), receipt of coronary angiography or PCI and cardiology care. For patients undergoing PCI, the association between ‘COVID-19 ACS’ status and mortality was estimated by constructing separate models adjusting all the procedural information reported in table 3 and in addition to the aforementioned confounders. The independent predictors of 30-day mortality were studied using multivariable logistic regression model. All tests were two-sided and statistical significance considered as P< 0.05. Statistical analyses were performed in Stata MP 16.0 College station, Texas, US via secure remote access on the NHS Digital servers hosting all the datasets.

## Results

Between 1^st^ March and 31^st^ May 2020, 12,958 patients were hospitalized with ACS in England, of which 517 (4.0%) were COVID-19 positive. There was a steady increase in the number of daily COVID-19 ACS hospitalizations during March, reaching a peak in the first week of April followed by a steady decline by the end of May (Figure 1). Higher proportions of daily COVID-19 ACS cases were admitted with a suspected NSTEMI compared to STEMI throughout the study period.

### Patient characteristics

Patients in the COVID-19 ACS group were older compared with the non COVID-19 ACS group (72.8 year vs 67.0 year), and a greater proportion were from Black, Asian and Ethnic minority origin (20.2% vs 12.8%), they had a lower body mass index (26.9 vs 28.2) and more likely to be hospitalized with NSTEMI (67.0% vs 62.0%). The COVID-19 ACS group also exhibited an increased incidence of in-hospital cardiac arrest (6.3% vs 3.0%), higher troponin levels, and were more likely to have presented in pulmonary oedema (9.0% vs 3.4%) or cardiogenic shock (9.6% vs 3.9%). They had a higher prevalence of heart failure (23.7% vs 13.4%), cerebrovascular disease (15.7% vs 8.0%), insulin treated diabetes (13.6% vs 7.5%) and hypertension (69.4% vs 58.3%).

Out of 12,958 patients, 6,864 (53.0%) underwent PCI and were successfully linked from MINAP into the BCIS registry (supplementary figure 1). COVID-19 ACS patients undergoing PCI were of similar age and had similar baseline characteristics to the overall ACS cohort (Table 2). The angiographic and procedural profiles of COVID-19 ACS patients (such as number of lesions attempted, vessels attempted, multivessel PCI, number of stents used, use of intracoronary imaging such as IVUS, OCT and pressure wire use) were similar to the non COVID-19 ACS cohort undergoing PCI. From the patients who didn’t receive PCI, the proportions of angiographically normal coronaries or surgical disease were similar in both non COVID-19 and COVID-19 ACS groups.

**Table 1:**
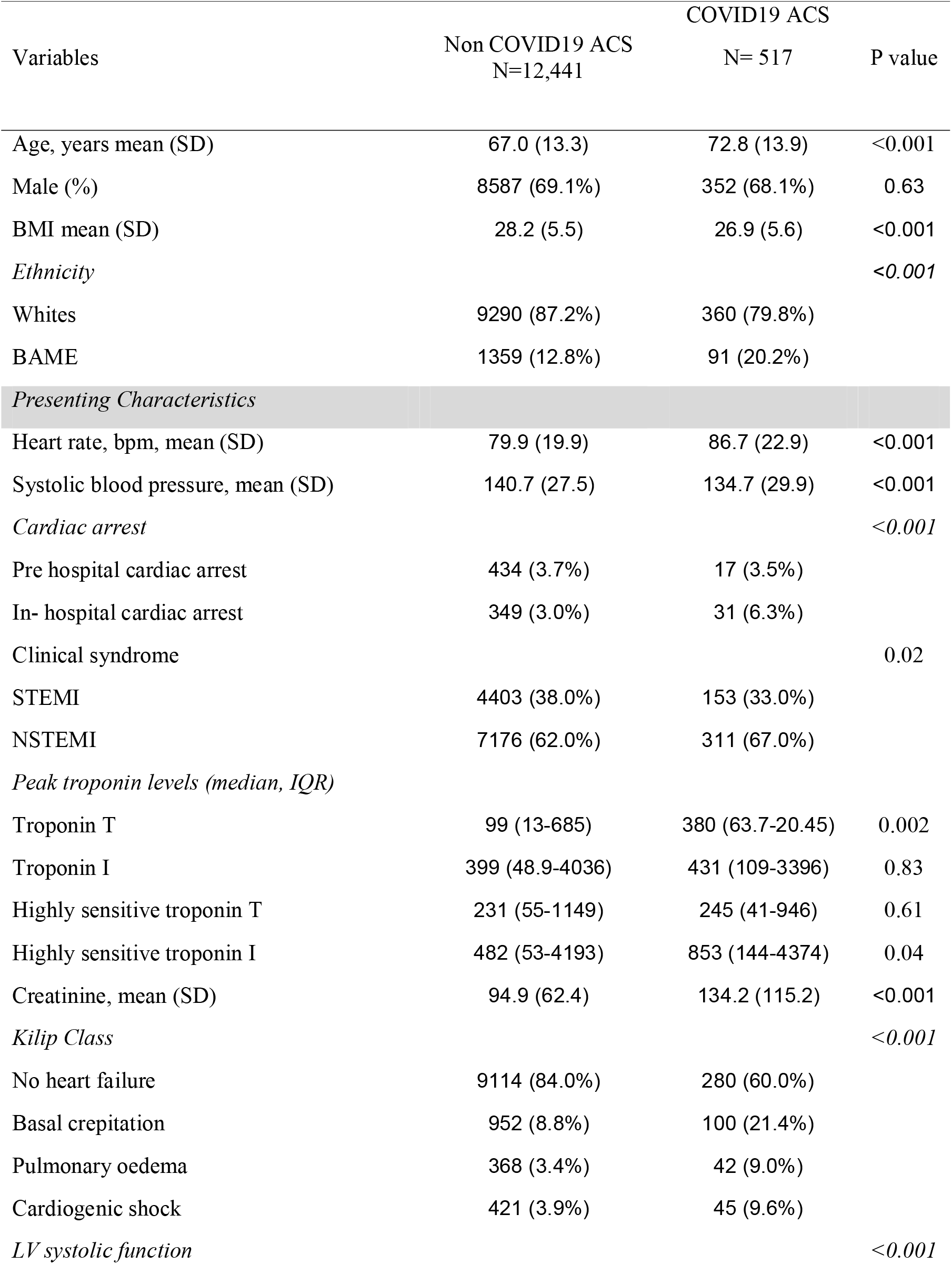
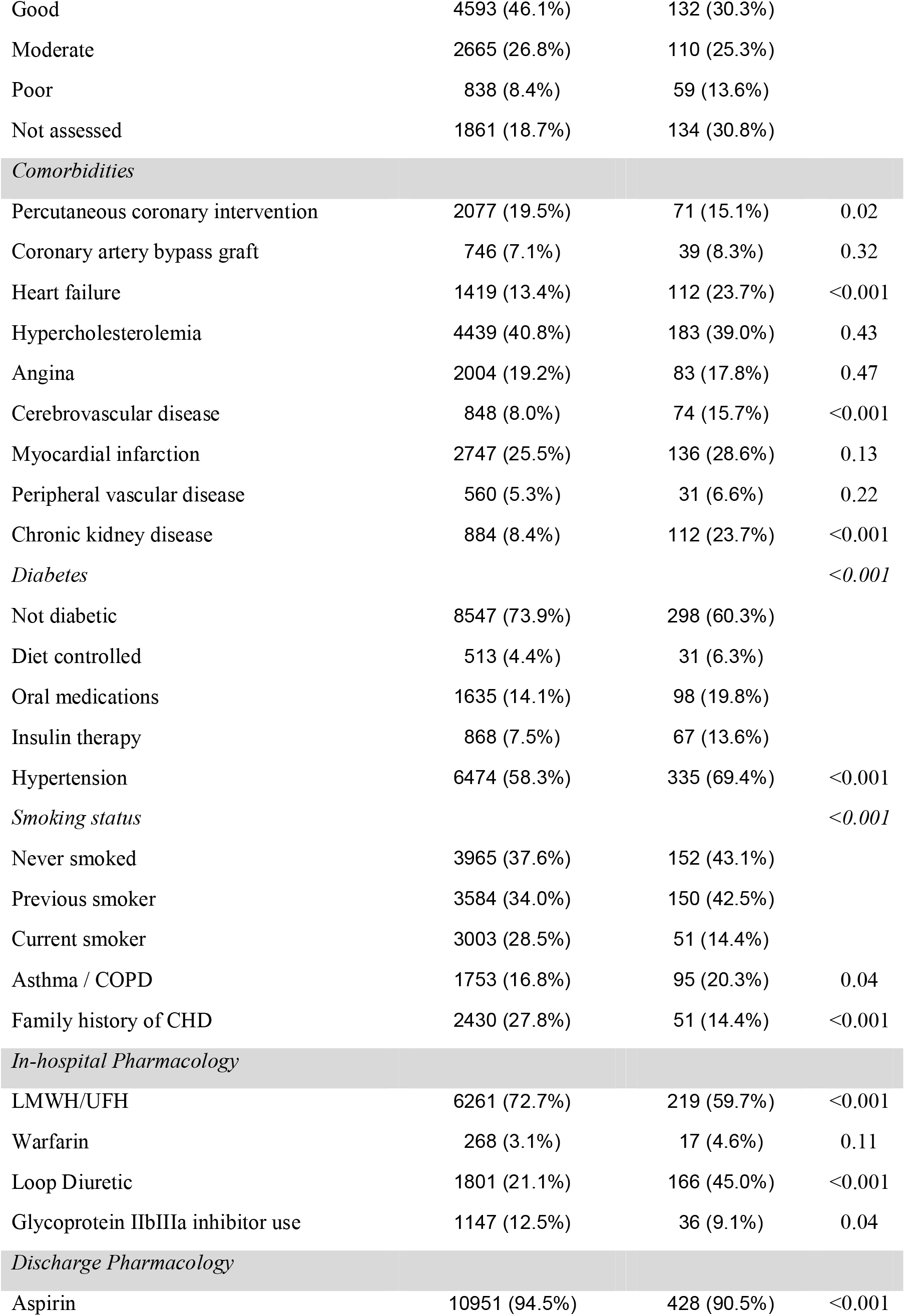
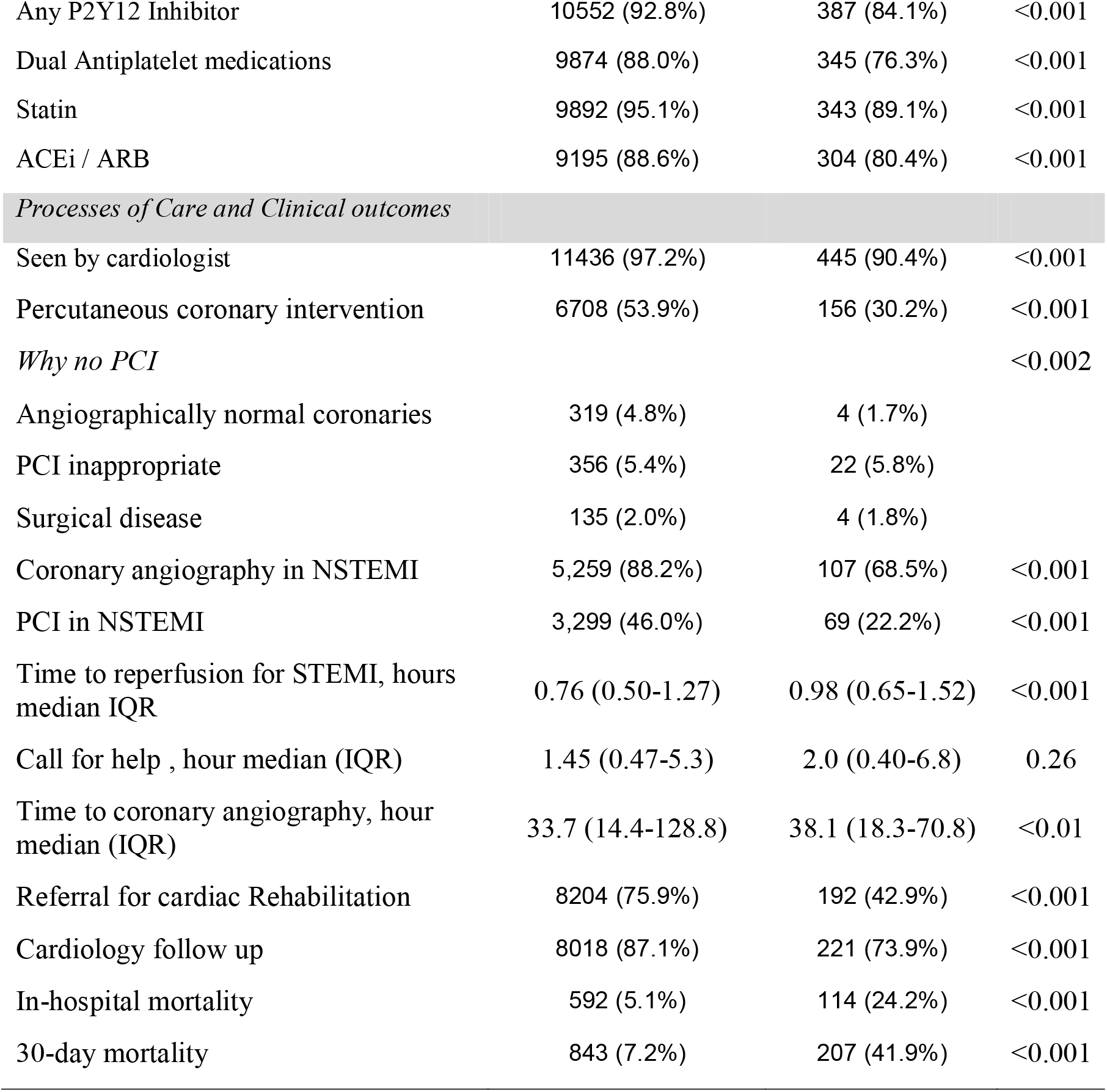
Baseline characteristics COVID19 ACS compared to non COVID19 ACS patients in the MINAP registry.

**Table 2:** Clinical characteristics of COVID19 ACS compared to non COVID19 ACS patients undergoing PCI in the BCIS registry.

| Variables | Non COVID19 ACS<br>N=6708 | COVID19 ACS<br>N= 156 | P value |
| --- | --- | --- | --- |
| Age, years mean (SD) | 64.4 (12.0) | 65.3 (12.5) | 0.38 |
| Male (%) | 5019 (74.9%) | 118 (75.6%) | 0.84 |
| BMI mean (SD) | 28.5 (5.2) | 27.9 (4.8) | 0.17 |
| *Ethnicity* |  |  | <0.001 |
| Whites | 5045 (85.5%) | 104 (72.7%) |  |
| BAME | 853 (14.5%) | 39 (27.3%) |  |
| *Presenting Characteristics* |  |  |  |
| Heart rate, bpm, mean (SD) | 78.1 (18.4) | 83.8 (20.9) | <0.001 |
| Systolic blood pressure, mean (SD) | 140.3 (27.1) | 137.1 (31.5) | 0.18 |
| *Cardiac arrest* |  |  | <0.001 |
| Pre hospital cardiac arrest | 274 (4.3%) | 9 (6.2%) |  |
| In- hospital cardiac arrest | 179 (2.8%) | 14 (9.7%) |  |
| Clinical syndrome |  |  | 0.21 |
| STEMI | 3394 (50.7%) | 87 (55.8%) |  |
| NSTEMI | 3299 (49.3%) | 69 (44.2%) |  |
| Creatinine, mean (SD) | 89.2 (53.0) | 114.6 (105.2) | <0.001 |
| *Kilip Class* |  |  | <0.001 |
| No heart failure | 5224 (87.8%) | 95 (66.0%) |  |
| Basal crepitation | 336 (5.6%) | 18 (12.5%) |  |
| Pulmonary oedema | 136 (2.3%) | 14 (9.7%) |  |
| Cardiogenic shock | 251 (4.2%) | 17 (11.8%) |  |
| *LV systolic function* |  |  | 0.21 |
| Good | 2750 (50.8%) | 58 (43.6%) |  |
| Moderate | 1704 (31.5%) | 50 (37.6%) |  |
| Poor | 421 (7.8%) | 14 (10.5%) |  |
| Not assessed | 539 (10.0%) | 11 (8.3%) |  |
| *Comorbidities* |  |  |  |
| Percutaneous coronary intervention | 1265 (22.0%) | 31 (21.8%) | 0.96 |
| Coronary artery bypass graft | 303 (5.4%) | 11 (7.9%) | 0.22 |
| Heart failure | 542 (9.6%) | 20 (14.6%) | 0.05 |
| Hypercholesterolemia | 2949 (49.5%) | 72 (51.1%) | 0.72 |
| Angina | 812 (14.6%) | 14 (10.4%) | 0.17 |
| Cerebrovascular disease | 361 (6.4%) | 15 (10.8%) | 0.03 |
| Myocardial infarction | 1412 (24.4%) | 42 (29.8%) | 0.14 |
| Peripheral vascular disease | 282 (5.0%) | 14 (10.1%) | 0.007 |
| Chronic kidney disease | 274 (4.9%) | 22 (16.1%) | <0.001 |
| *Diabetes* |  |  | <0.001 |
| Not diabetic | 4716 (75.5%) | 91 (60.7%) |  |
| Diet controlled | 274 (4.4%) | 6 (4.0%) |  |
| Oral medications | 872 (14.0%) | 33 (22.0%) |  |
| Insulin therapy | 384 (6.1%) | 20 (13.3%) |  |
| Hypertension | 3678 (60.2%) | 104 (71.7%) | 0.005 |
| *Smoking status* |  |  | 0.02 |
| Never smoked | 2462 (38.3%) | 66 (46.2%) |  |
| Previous smoker | 2040 (31.7%) | 49 (34.3%) |  |
| Current smoker | 1934 (30.0%) | 28 (19.6%) |  |
| Asthma / COPD | 765 (13.7%) | 18 (13.3%) | 0.90 |
| Family history of CHD | 1542 (31.8%) | 28 (26.2%) | 0.21 |
| *In-hospital Pharmacology* |  |  |  |
| Low molecular weight heparin | 1867 (43.2%) | 39 (42.4%) | 0.88 |
| Unfractionated heparin | 2092 (47.2%) | 34 (36.2%) | 0.03 |
| Warfarin | 98 (2.2%) | 5 (5.4%) | 0.04 |
| Loop Diuretic | 676 (15.6%) | 31 (33.7%) | <0.001 |
| Glycoprotein IIb/IIIa inhibitor use | 1015 (20.7%) | 32 (27.6%) | 0.07 |
| *Discharge Pharmacology* |  |  |  |
| Aspirin | 6226 (98.2%) | 138 (97.9%) | 0.76 |
| Any P2Y12 Inhibitor | 6013 (97.3%) | 121 (92.4%) | <0.001 |
| Dual Antiplatelet medications | 5827 (95.6%) | 116 (89.9%) | 0.002 |
| Statin | 5645 (97.6%) | 107 (95.5%) | 0.17 |
| ACEi / ARB | 5391 (93.1%) | 110 (94.0%) | 0.70 |
| *Processes of Care and Clinical outcomes* |  |  |  |
| Referral for cardiac Rehabilitation | 5068 (86.3%) | 84 (64.1%) | <0.001 |
| Cardiology follow up | 4662 (93.3%) | 87 (89.7%) | 0.16 |
| In-hospital mortality | 180 (2.8%) | 19 (14.0%) | <0.001 |
| 30-day mortality | 260 (4.1%) | 44 (30.8%) | <0.001 |

**Table 3:**
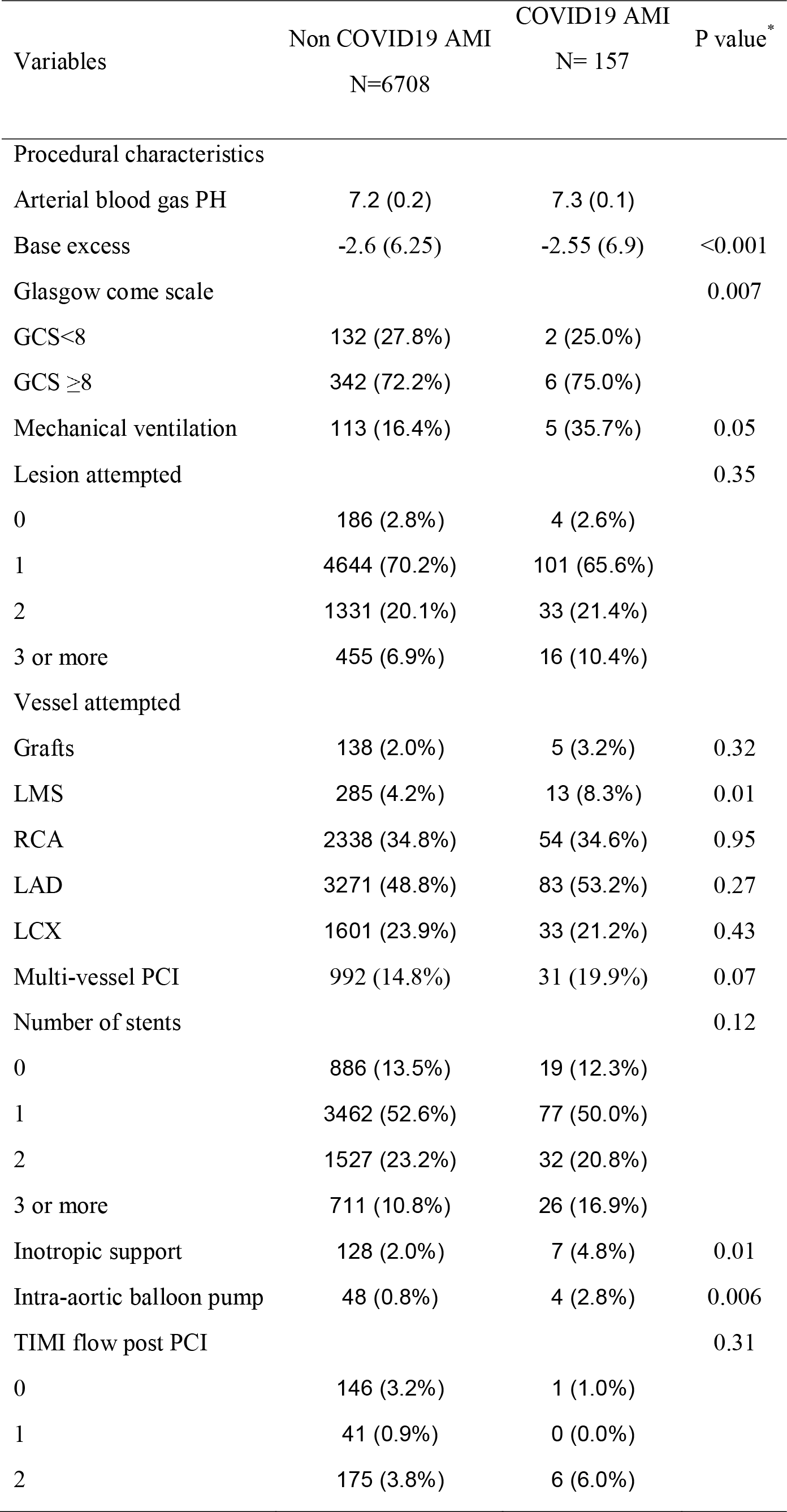
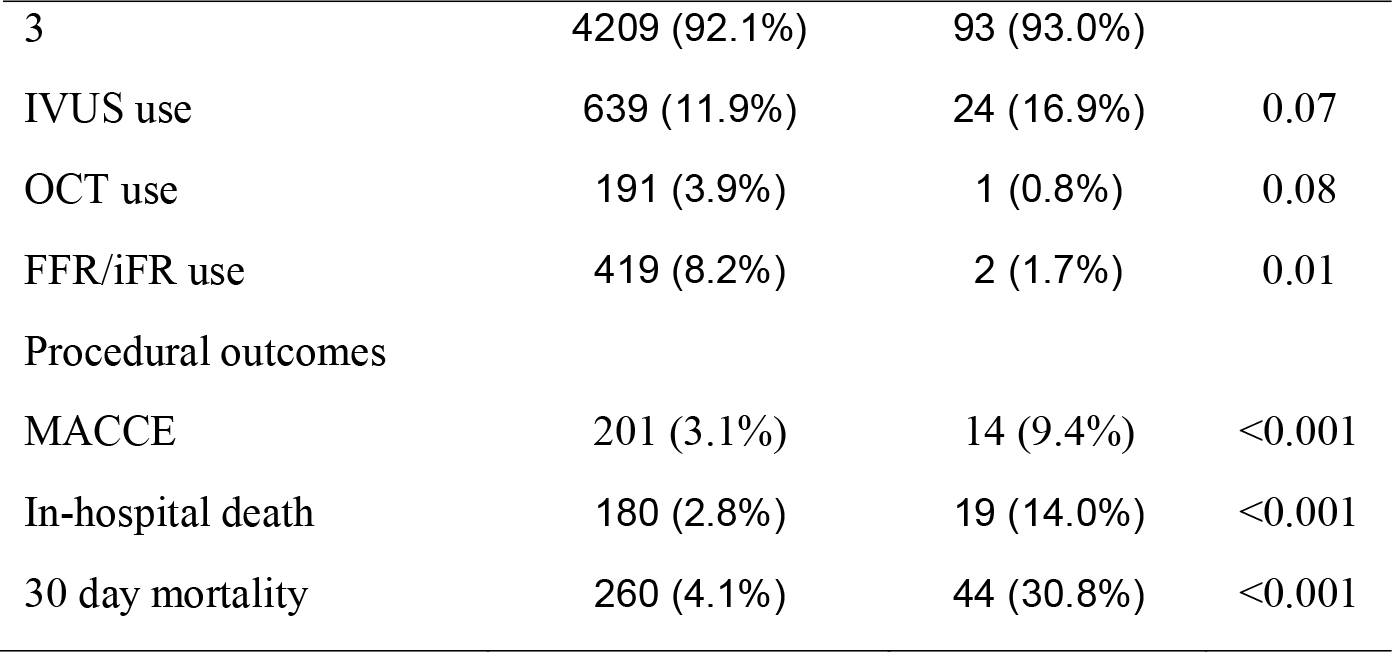
Angiographic characteristics and procedural outcomes of COVID19 ACS patients compared to non COVID19 ACS patients undergoing PCI in the BCIS registry.

### Discharge medication and Guidelines-indicated care

Patients in the COVID-19 ACS group were less likely to receive optimal secondary prevention medication such as aspirin (90.5% vs 94.5%), dual anti-platelet medications (76.3% vs 88.0%), statin (89.1% vs 95.1%) and ACE/ARB (80.4% vs 88.6%) therapy compared with the non COVID-19 ACS group. Overall, only one third (30.2% vs 53.9%) of COVID-19 ACS patients received PCI compared to non COVID-19 ACS patients. COVID-19 NSTEMI patients were also less likely to undergo invasive coronary angiography (67.7% vs 81.0%) and PCI (22.2% vs 46.0%) compared to the non COVID-19 NSTEMI cohort.

The median time from in-hospital arrival to reperfusion for patients presenting with STEMI was 13.2 minutes longer in the COVID-19 STEMI group compared with the non COVID-19 STEMI group. COVID-19 NSTEMI group also experienced delays in time to coronary angiography (38.1 hours vs 33.7 hours) compared with non COVID-19 NSTEMI group. Less than half of the COVID-19 ACS group were referred to the cardiac rehabilitation programme (42.9% vs 75.9%) or had cardiology follow-up arranged following discharge from the hospital (73.9% vs 87.1%).

### Clinical outcomes and predictors of mortality

In the MINAP cohort, the COVID-19 ACS group had higher in-hospital (24.2% vs 5.1%) and 30-day mortality rates (41.9% vs 7.2%) compared with the non COVID-19 ACS cohort. After adjustment for baseline differences in demographics, presenting characteristics, comorbidities and pharmacology, the hierarchical multi-level logistic regression model showed a significantly higher risk of in-hospital (adjusted odds ratio 3.27 95%CI 2.41-4.42) and 30-day mortality (adjusted odd ratio 6.53 95%CI 5.12-8.36) in the COVID-19 ACS patients compared with non COVID-19 ACS patients (Figure 2). In the subgroup analysis stratified according to type of ACS, although short term mortality was similar between the NSTEMI and STEMI groups, the NSTEMI group had higher mortality rates (adjusted odds ratio 8.45 95%CI 6.03-11.83) at 30-days. Similar, higher mortality rates were observed in COVID-19 ACS patients undergoing PCI compared with non COVID-19 ACS patients undergoing PCI. The multilevel regression model showed that increasing age per year, severe LVSD, in-hospital cardiac arrest, peak troponin concentration levels, renal dysfunction and use of ACE/ARB on discharge were strong independent risk factors for 30-day mortality in the COVID-19 ACS group (Figure 3).

**Figure 2:**
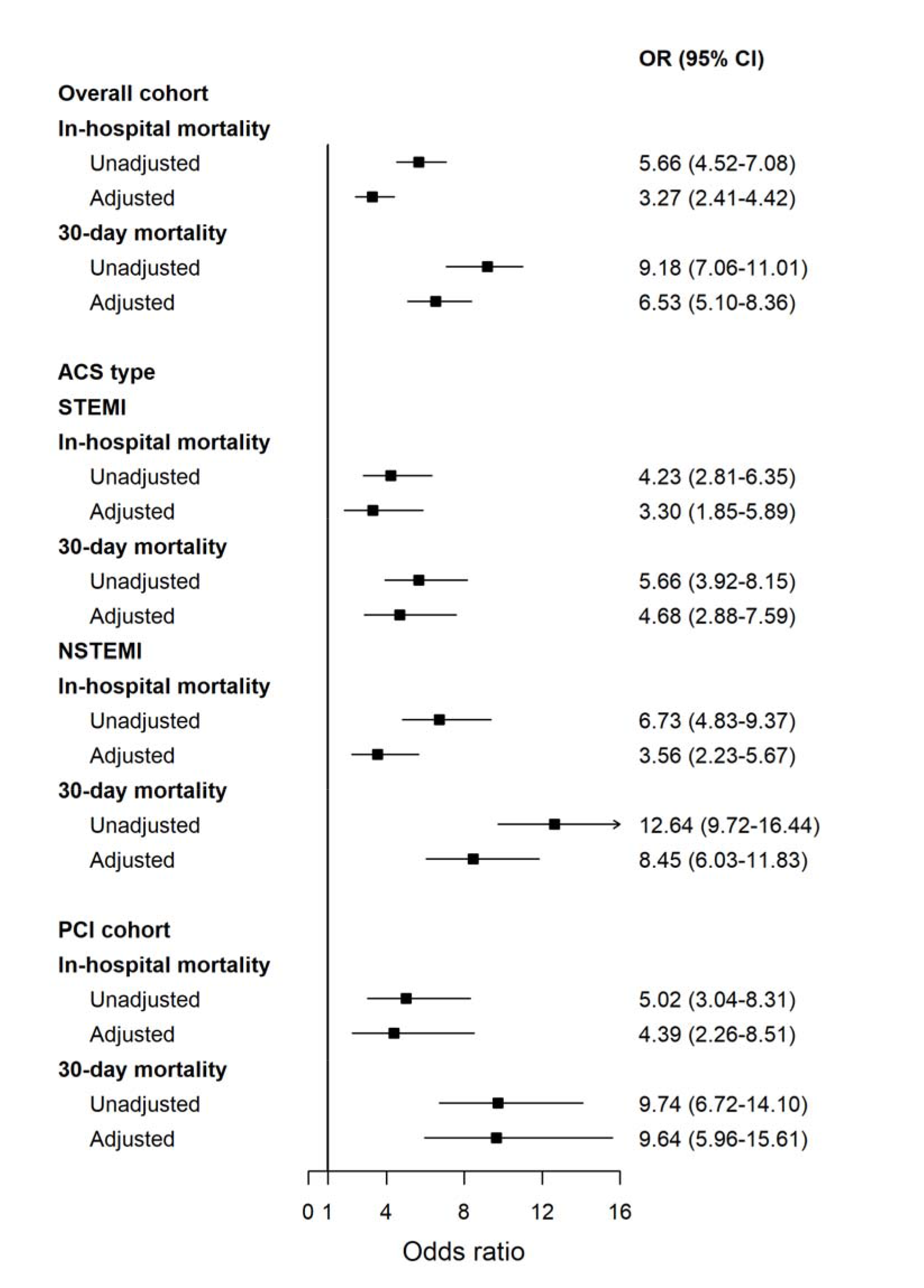
In hospital outcomes and 30-day mortality of COVID19 ACS patient compared to non COVID19 ACS patients

**Figure 3:**
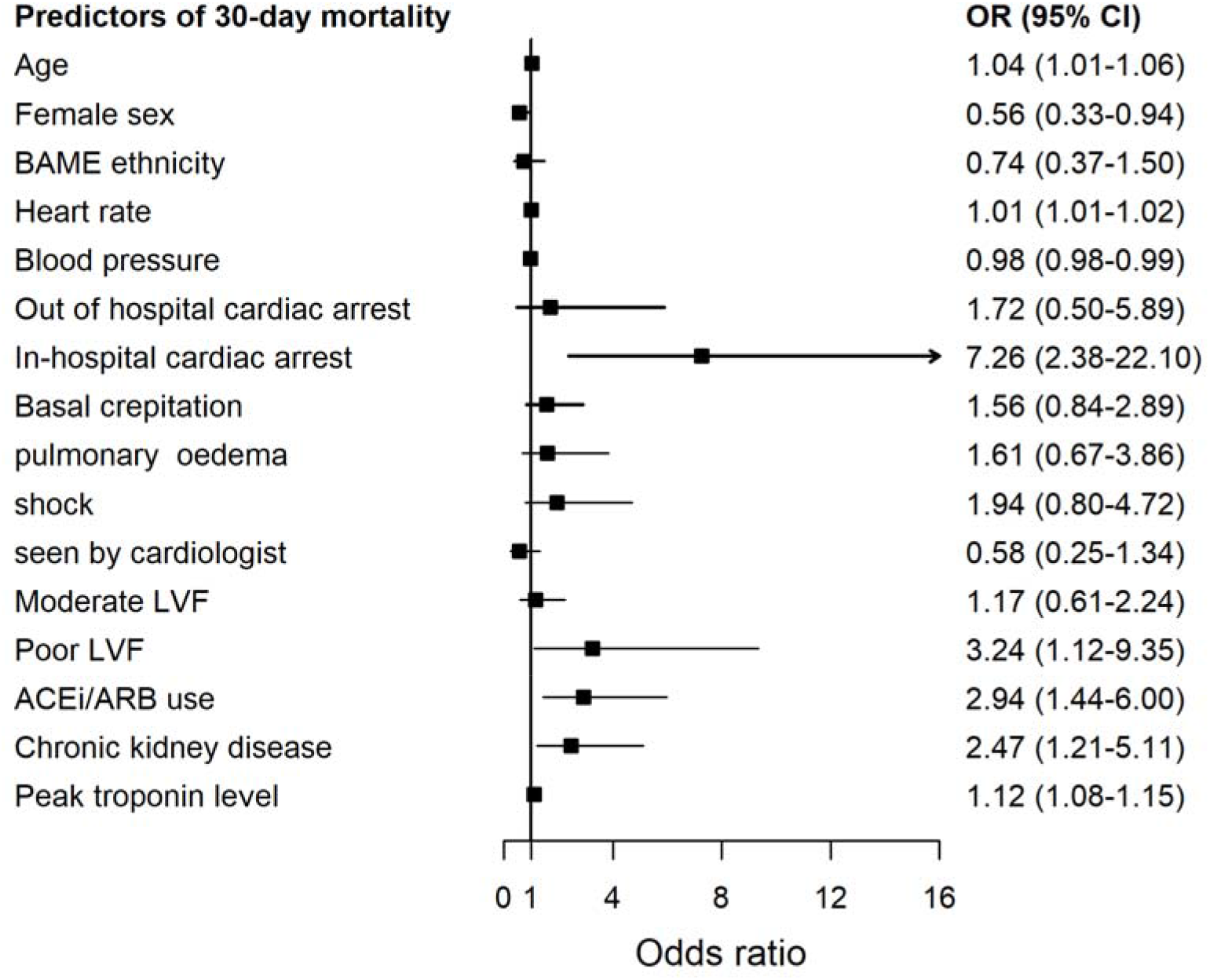
Independent predictors of 30-day mortality in COVID positive AMI patients

## Discussion

This multisource, national report of contemporary data from all ACS related hospitalizations in England during the COVID-19 period provides detailed information about the risk profile, clinical care and outcomes of ACS patients with COVID-19 infection. Specifically, COVID-19 ACS group had over 6-fold increase in mortality within 30 days compared with non COVID-19 ACS and were less frequently prescribed appropriate secondary prevention medications. They were older, more likely to be of BAME origin, had more comorbid features and exhibited high risk presenting characteristics such as higher Killip class, troponin concentrations, creatinine and evidence of LV systolic dysfunction.

A significant knowledge gap exists in the literature about incidence and profile of ACS patients with concomitant COVID-19 diagnosis, and their associated clinical outcomes. We report that COVID-19 infection in patients presenting with ACS is prevalent at a low level in a national cohort of patients. The higher prevalence of pulmonary oedema and shock at presentation, together with the higher troponin concentrations levels in the COVID-19 ACS cohort are suggestive of increased myocardial injury in this group corroborating findings from China and US cohorts^19,20^. However, contrary to earlier small case series from New York and Italy^8,9,21^, the angiographic characteristics of COVID-19 ACS patients were not different from the non COVID-19 ACS patients, with a very small prevalence of non-obstructive coronary disease and similar procedural success.

Whilst previous studies have reported the incidence on myocardial injury based on cardiac biomarker levels in patients hospitalized with a confirmed COVID-19 diagnosis^19,20^, none reported on management and outcomes of ACS patients with concomitant COVID-19 diagnosis in a national, unselected cohort of suspected type 1 myocardial infarction. The management of ACS patients during the COVID-19 pandemic is based on expert clinical guidance which lacks consensus on the optimal treatment strategies for patients presenting with ACS during the COVID-19 outbreak^22–25^. The Chinese Cardiac Society consensus statement proposed medical management for the majority of patients presenting with non-ST-elevation myocardial infarction (NSTEMI), and thrombolysis in those presenting with STEMI during the COVID-19 pandemic^22^. In contrast, the North America and Canadian guidelines recommended use of thrombolysis as an alternative to primary PCI for patients with STEMI particularly where PCI services are restricted due to COVID-19^23,25^. However, a joint statement from British Cardiovascular Society, BCIS and NHS England in the UK recommended that primary PCI should remain the default treatment for all STEMI, except in unusual circumstances^26^. In keeping with this national recommendation, we observed almost negligible use of thrombolysis for STEMI in the UK during the acute phase of the COVID-19 pandemic. By contrast, there were significant differences in the care of COVID-19 ACS patients in that only a third of COVID-19 ACS patients received PCI, they experienced greater delays in reperfusion treatment or an invasive strategy and had a significantly lower uptake of secondary prevention medications on discharge. Given that COVID-19 ACS patients presented with more complex, high risk features, there is an urgent need to develop effective treatment pathways to align their care with guideline recommendations^27,28^. For patients hospitalized with suspected ACS, especially with elevated cardiac troponin minor ECG changes, an immediate COVID-19 testing should be advocated in the current climate.

In the outcomes analysis, COVID-19 ACS patients had a poor prognosis compared with the non COVID-19 ACS patients. One of four patients died in hospital with an over six-fold higher 30-day mortality in the overall cohort as well as those undergoing PCI. Although we didn’t have information regarding the presenting COVID-19 symptoms or respiratory complications in this cohort, we noted significantly higher levels of peak troponin, creatinine, a lower presenting blood pressure and tachycardia in this cohort. Troponin elevation three times the upper reference limit is known to be associated with worse in-hospital outcomes in COVID-19 patients^19,20^. It remains unclear whether the rise in cardiac biomarkers is related to viral myocarditis^5,6^ or plaque rupture secondary to virus-induced inflammatory response^29–31^ or a type 1 AMI. In this cohort of suspected type 1 myocardial infarction, there was a low prevalence of non-obstructive coronary disease and the angiographic characteristics and procedural management of the COVID-19 ACS were similar to the non COVID-19 ACS group. We observed higher rates of COVID19 infection and subsequent 30-day morality in NSTEMI compared with STEMI. Patients presenting with NSTEMI are generally older, more comorbid and less likely to present early during the COVID-19 pandemic which in conjunction with increased infection rates may be responsible for their poor outcomes.

The COVID-19 ACS cohort were also less frequently prescribed low molecular weight or unfractionated heparin and statins. Patients with acute COVID-19 are known to be at increased risk of thromboembolic complications and are therefore likely to benefit from appropriate anticoagulation to reduce thrombus burden and statins to stabilize plaque. In the risk factor analysis, elevated creatinine, peak troponin, heart rate, left ventricular systolic dysfunction and use of ACE inhibitor or ARBs were strong independent risk factors for 30-day mortality in this report. Although recent data have negated earlier concerns of an increased risk of adverse outcomes in COVID-19 patients with use of ACE inhibitors or ARBs, further research is required to establish the role of these medications in patients presenting with COVID-19 ACS^32,33^. Patients hospitalized with ACS and concomitant COVID-19 infection may also be focus of specific anti-viral drug trials or immunosuppression therapy to establish the benefit of these therapies in this high risk cohort^7^.

The interrogation of multisource, multihospital live data from England has given insight in to the presenting characteristics, risk profile, treatment strategies and 30-day mortality in an unselected cohort of patients hospitalized with COVID-19 ACS. However, there are some study limitations which must be kept in mind whilst interpreting these findings. First, the COVID-19 diagnosis was based on the ICD-10 codes and so it is unclear whether the diagnosis at the hospital was made on clinical grounds or using a formal polymerase chain reaction (PCR) test, and the timing of the tests leading to the diagnosis is unclear. Moreover, other suspected ACS patients were not routinely tested at the start of the pandemic and we cannot say how many might have been carrying the virus, although the lack of a COVID-19 code suggests they did not have a clinical syndrome related to the virus. Second, information around COVID-19 symptoms, their duration and other organ involvement is not captured in the datasets used in this analysis, and hence it difficult to ascertain whether patients had COVID-19 symptoms followed by an ACS or vice versa, and whether COVID-19 infection occurred in the hospital or the community. Third, whilst the MINAP registry is the UK national ACS registry for suspected type1 AMI, we cannot rule out misclassification bias from misdiagnosis of myocarditis or type 2 AMI, although only a small proportion of cases that underwent angiography had non-obstructive coronary artery disease.

## Conclusion

COVID-19 infection was present in 4% of patients hospitalized with a suspected ACS in England during the study period. It was associated with worst outcomes independent of the type of ACS, however higher 30-day mortality was observed in NSTEMI group compared with STEMI group. Elderly, comorbid and BAME patients with underlying cardiovascular disease are more likely to acquire COVID-19 infection and often present with unfavourable presenting characteristics. These findings highlight the need to develop strategies for an efficient and equitable care for COVID ACS patients.

## Data Availability

Not applicable

## Acknowledgments

The authors acknowledge Chris Roebuck, Tom Denwood, Tony Burton and Courtney Stephenson and data support staff at NHS Digital for providing and creating the secure environment for data hosting and for analytical support, and Anil Gunesh and Julian Hains from the National Institute of Cardiovascular Outcomes Research for data transfer into the secure environment. This work uses data provided by patients and collected by the NHS as part of their care and support and authors would like to thank all for their contributions.

